# The performance of *in silico* prediction tools for variant curation in a panel of cancer genes

**DOI:** 10.1101/2025.07.29.25331316

**Authors:** Niles Nelson, Aram Niaz, Kirsten Fairfax, Tracy M. Bryan, Sionne Lucas, Joanne Dickinson

## Abstract

Rare single base pair changes in genes are an important cause of disease, as they can reside in key regions of the gene influencing biological function by impacting the protein conformation and protein interactions. Generation of the necessary experimental evidence to define the outcome of the presence of these gene variants is time consuming and costly. These challenges have led to the development of a plethora of *in silico* prediction tools. These tools frequently use similar sources of information and are trained on overlapping multi-gene ‘truth’ datasets. However, frequently there has been no quantitative validation of the performance of these *in silico* tools for individual genes. Here we have applied the ClinGen Sequence Variant Interpretation Working Group’s recommended *in silico* score thresholds to a set of predisposition gene variants with established pathogenicity/benignity. Of the genes assessed (*BRCA1, BRCA2, TP53, TERT* and *ATM*), *in silico* tool predictions showed inferior sensitivity (<65%) for pathogenic TERT variants and inferior sensitivity (≤81%) for benign TP53 variants. This validation study highlights *in silico* tool performance can be gene-specific and is dependent on the ‘training set’ on which the algorithm is built. Where there are sufficient numbers of established benign and pathogenic missense variants based on clinical and functional evidence, the use of *in silico* tool scores should be validated for individual genes. For genes where this is not possible and gene-agnostic *in silico* score cut offs are used, consideration of missense variant-protein structural impact relationships is suggested.

## Introduction

The interpretation of missense variants detected in cancer predisposition genes remains a significant challenge both for those in the clinical and research settings. Pathogenic variants can be ultra-rare or even private to individual families, making enrichment in cases difficult to establish^1^. Phenotypes are often late-onset, incompletely penetrant and non-pathognomonic, resulting in limited utility of segregation evaluation^1,2^. Where affected family members are identified, it is also likely to be posthumously, therefore hampering segregation testing. While functional genomic assays are available to permit assessment of pathogenicity/benignity for some genes (*TP53^3,4^, BRCA1^5,6^, BRCA2^7^* and *TERT*^8^), for many genes there remains a dearth of assays addressing function. The significant cost in terms of assay development, the labour-intensiveness, and the low throughput associated with functional genomic studies also impacts the practical application for variant curation. Use of the American College of Medical Genetics and Association for Molecular Pathology (ACMG/AMP) criteria to permit formal curation of the variants to inform clinical decisions is therefore problematic as for the vast majority of rare missense variants identified, the table of criteria cannot be applied^2,9^.

*In silico* tools provide an opportunity to assist with the interpretation of the likely pathogenicity of rare variants both in the research and clinical settings. The ACMG/AMP criteria PP3 and BP4 were established to assist with the use of *in silico* tools for diagnostic curation^9^. The PP3 criterion is used to apply *in silico* data supporting pathogenicity of a variant. The BP4 criterion is used conversely to apply *in silico* data supporting benignity of a variant. As gene-specific curation guidelines have developed, variant curation working groups have provided gene-specific recommendations regarding the use of *in silico* prediction tools^1,10^. While, ideally *in silico* tools should be validated for individual genes, in reality, this is challenging. For many genes there are insufficient established pathogenic and benign missense variants for validation of these tools and we must rely on score thresholds determined by algorithm training on aggregated data from multi-gene ‘truth sets”.

Evaluation of the performance of *in silico* tools has shown the potential to improve the curation of missense variants by adjusting the score thresholds to allow higher confidence in pathogenic or benign predictions^11,12^. This study aims to assess the performance of these gene-agnostic recommendations when applied to individual genes harbouring rare variants contributing to cancer risk. In addition to evaluating predictive performance of the *in silico* tools, REVEL^13^, MutPred2^14^, Bayesdel (no AF)^15^, VEST4^16^ and CADD^17,18^, we examined the impact of the recommended thresholds on predictive performance and whether the addition of an *in silico* prediction tool evaluating the impact of the variant on functionality of the splice site (Splice AI)^19^ where appropriate, or protein structure, MISCAST^20^ can improve prediction accuracy.

## Material and Methods

To examine the performance of *in silico* tools in predicting pathogenicity we examined the thresholds imposed to interpret the predictions scores generated by these tools (table 1)^11^. Bayesian statistical analyses have been used to determine associated target positive likelihood ratios (PLRs)^21,22^. It is estimated that in the clinical setting, diagnostic genomic testing will reveal a likely pathogenic variant in approximately 10% of the variants identified. This underlying estimate of prior probability (0.1) has been used to inform application of the ACMG/AMP criteria to deliver curated scores for pathogenicity. This has equated to PLRs (defined as an OddsPaths) of greater than 2.1, 4.3, 18.7 and 350 to achieve the thresholds of “supporting”, “moderate”, “strong” and “very strong strength levels” respectively^22^. The final posterior probability associated with this approach to curation is estimated to >0.9 for a likely pathogenic curation and >0.99 for a pathogenic curation. The inverse of this applies to benign and likely benign curations.

**Table 1.**
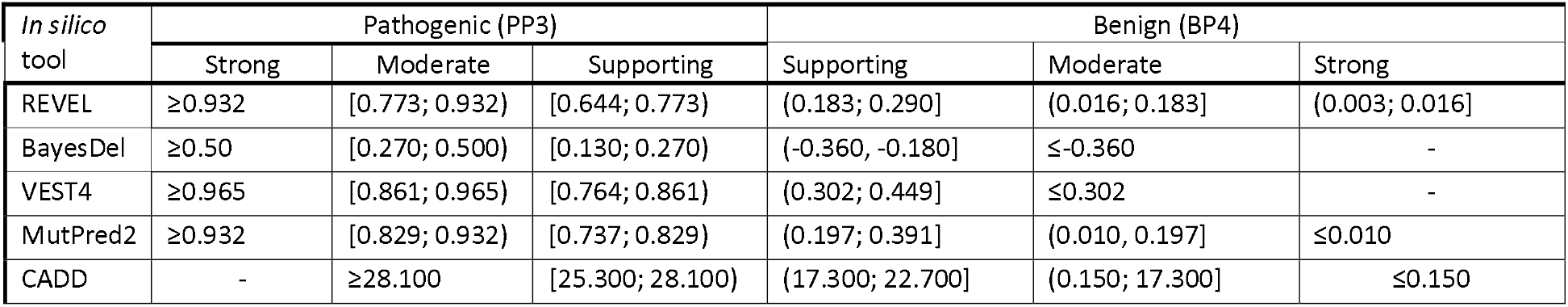
*In silico* tool score cut-offs used in this study (derived from Pejaver et al., 2022).

### Selection of *in silico* tools

The following tools were selected for analysis in this study: REVEL^13^, MutPred2^14^, Bayesdel (no AF)^15^, VEST4^16^ and CADD^17,18^. In silico tool selection was based on the breadth of evidence strength determined by Pejaver et al. 2022. Tools were prioritized that potentially provided up to a strong level of evidence to meet the ACMG criteria for PP3 or BP4.

REVEL, a random forest classifier, integrates functional impact prediction tool results (MutationAssessor^23^, MutationTaster^24^, PolyPhen-2^25^, LRT^26^ and SIFT^27^), conservation score tool results (GERP++^28^, PhyloP^29^ and SiPhy^30^), predictions from other meta-predictors (FATHMMv2.3^31^, PROVEAN^32^ and VEST3^33^) as well as protein domain and population allele frequency data for a given missense variant to predict pathogenicity^13^. MutPred2 utilises a deep neural network trained on curation data (ClinVar^34^, HGMD^35^), interspecies pairwise alignments, population allele frequency data (ExAC^36^) and protein structural and functional data (UniProt^37^ and the protein data bank^38^). BayesDel(no AF) is also an ensemble tool based on a native Bayes classifier trained on curation data (ClinVar, HGMD), the same functional impact and conservation tools utilised for REVEL. Variants were also annotated with FATHMM predictions. Population allele frequency data was intentionally not included to assist with rare variant interpretation. VEST4, also a random forest classifier, utilises disease (HGMD) and population (ESP6500^39^) data and SNVBox^40^ annotations to generate predictions. While similar conservation and functional data usage is similar (Grantham, Polyphen, SIFT and others) CADD is unique amongst these tools, in that the underlying algorithm was not trained on training sets of known disease-causing gene variants with established non-VUS classifications. CADD also integrates some splice prediction into the scoring algorithm^15,17^. AlphaMissense is an additional *in silico* tool that has been developed subsequent to the above described analysis, however, the initial validation analysis is suggestive it may outperform many these other established tools^41^. The variants in the TERT_3 and TERT_1 datasets were annotated with AlphaMissense scores (Table S6, Table S7) as a comparison to the tools reviewed by Pejaver et al., 2022.

SpliceAI^19^ was also included in the analysis as it is commonly used in combination with other *in silico* tools for ClinGen VCEP curation guidelines^1^, with the rationale that novel splice sites may be generated by single nucleotide variants in coding regions of genes. Cut-off thresholds were determined based on current usage in guidelines^2,42,43^. Briefly, based on receiver operator curves generated using a truth set of 201 non-canonical splice variants, SpliceAI thresholds of >0.38 and <0.2 were associated with 90% sensitivity and 90% specificity for pathogenicity and benignity respectively^2^.

### MISCAST to predict pathogenicity using protein structural impact prediction

*In silico* tool algorithms often incorporate predictions regarding the structural impact of missense variants on the translated protein. As it is not possible to determine how these predictions are incorporated into *in silico* algorithms, the influence of these predictions on an *in silico* score for a given missense variant is difficult to determine. MISCAST^20^ is an *in silico* tool that incorporates only structural impact predictions for missense variants which can give insight into the impact of these predictions on the overall *in silico* score for a given missense variant. The algorithm used to generate MISCAST predictions (P3DFi scores) was trained using similar gene variant reference sets as many of the commonly used *in silico* tools^13-15^. To develop the MISCAST algorithm, variants from 1,330 disease causing genes were extracted from the ClinVar and the HGMD databases and compared to gene variants detected in healthy population controls (data extracted from GnomAD) to identify protein structural impact features that are enriched in disease causing variants^20^. To identify if MISCAST predictions could predict pathogenicity for the gene variants included in this present study, variant summary reports were downloaded from MISCAST^20^ for *TERT, TP53, BRCA1, BRCA2* and *ATM* and annotated with the benign and pathogenic classifications from the truth sets discussed below. The proportion of pathogenic and benign variants were determined for each “all protein classes” P3DFi score as described previously^20^. The benign variant proportion was subtracted from the pathogenic variant proportion for the P3DFi scores <-2, -2, -1, 0, 1, 2, and >2 respectively. Result visualisation was generated using ggplot2; https://ggplot2.tidyverse.org. Code is available on request.

### Generation of ‘truth sets’ of missense variants for rule evaluation

In order to evaluate the positive predictive value and negative predictive value of an *in silico* score cut off, it must be tested on a set of known pathogenic and known benign variants; a truth set. For many cancer predisposition genes, there are insufficient numbers of variants with definitive pathogenic or benign classifications to create a truth set large enough for statistical analysis. Using the bootLR R package^44^ (see below) for an *in silico* tool that was 97% sensitive and 97% specific, a truth set would need at least 30 pathogenic and 31 benign variants to be sufficiently powered to assess a moderate strength level target (PLR > 4.3). Therefore the *TP53, BRCA1, BRCA2, ATM* and *TERT* genes were selected as their exists greater than 61 missense variants with benign or pathogenic classifications available for each gene on public variant curation databases. The ClinGen Evidence Repository of variants curated by variant curation expert panels was also used as a source of established benign and pathogenic variants^45,46^. For the purposes of this study, pathogenic and likely pathogenic variants were grouped together and considered as pathogenic and similarly, benign and likely benign variants were both considered benign. A truth set was defined as a set of variants that have not been classified as a variant of uncertain significance (VUS) as determined by the curation criteria applied in the relevant source database.

#### TP53

Two *TP53* truth sets were used in this analysis. The first *TP53* missense variant truth set was obtained from Fortuno et al, 2018^47^. Pathogenicity and benignity were determined based on variant allele frequency in gnomAD, dominant negative effect and results from transactivation assays. Of the 318 variants that were available for analysis, 248 were pathogenic and 70 were benign. REVEL scores and BayesDel (no allele frequency) scores were obtained from the publication^47^. VEST4, CADD and MutPred2 scores were obtained from the dbNSFP (version 4.2) database of *in silico* tools on the 11^th^ August 2022. (Table S1)^48^. Following variant data upload, output of *in silico* results for 269 variants (248 pathogenic and 21 benign) was available for further analysis. The second truth set (TP53_2) used was derived from the TP53 variants included in the ClinGen Evidence Repository truth set discussed below (Table S2)^45,46^. Pathogenic variants were included if ACMG/AMP criteria for likely pathogenic or pathogenic were met without the use of PP3. Benign variants were included if at least one benign strong level criterion was met and no conflicting pathogenic moderate, strong or very strong criterion was met.

#### BRCA1 and BRCA2

*BRCA1* and *BRCA2* missense variants were obtained on the 16^th^ of July 2022 from the *BRCA1* and *BRCA2* Mutation Databases (University of Utah). Variant classification methods are described in the associated publication^49^. Briefly, gene-specific PLRs were estimated for co-occurrence with known deleterious variants, phenotypic and segregation data. Variants with combined odds of 1:100 and 1:1000 were considered likely pathogenic and pathogenic respectively^50^. For the*BRCA1* gene, 44 pathogenic and 267 benign missense variants were available for assessment. Following upload to the dbNSFP (version 4.2) database on the 11^th^ August 2022, *in silico* output (REVEL, VEST4, MutPred2, BayesDel (no allele frequency) and CADD) results for 43 pathogenic and 256 benign variants were obtained. *In silico* results were available for all 20 pathogenic and 286 benign *BRCA2* missense variants that were extracted (Tables S3, S4).

#### ATM

*ATM* missense variants were obtained from the Lieden open variation database (LOVD3) on the 16^th^ of July 2022^51^. Variants, as well as associated clinical and published data are submitted to LOVD3 from users at institutions with local LOVD3 instances. Variant curations (clinical classifications) are determined by the assigned LOVD3 curator. 40 pathogenic and 35 benign variants were extracted. In silico results were obtained for 36 and 33 variants respectively (Table S5).

#### TERT

The TERT_1 dataset comprises missense variants meeting ACMG/AMP criteria for pathogenicity or benignity without the use of the PP3/BP4 criterion. This dataset was derived from a comprehensive review and curation of all known *TERT* variants reported in the literature as well as the ClinVar and GnomAD databases. Details regarding the application of ACMG/AMP guidelines are described elsewhere^8^. Thirty-two variants with a pathogenic classification and only 6 variants with a benign classification were identified (Table S6). As reported previously, only a small minority of missense variants reported for *TERT* can be established as pathogenic or benign using current ACMG/AMP guidelines^8^ thus this dataset was underpowered to assess PLRs for PP3/BP4 at moderate strength levels. Therefore, sensitivity analysis was also undertaken using two related truth sets; TERT_2 (Table S6) and TERT_3 (Table S7). Details regarding these truth sets are included in the supplemental tables.

#### ClinGen data

Missense variants were also extracted for each of the genes from the ClinGen Evidence Repository on the 16^th^ of July 2022^46^. The ClinGen Evidence Repository provides detailed and standardized variant curation information for all included variants. 738 pathogenic and 361 benign missense variants from 50 genes were available for extraction (Table S8). *In silico* results were obtained from 721 and 352 variants respectively. A second truth set of ClinGen variants was also assessed to attempt to control for variant classifications dependent on use of *in silico* tool results. In this truth set pathogenic variants were considered as variants that met ACMG/AMP guidelines for likely pathogenic or pathogenic without the use of PP3. Benign variants were considered as variants where a benign code could be applied at a strength level of strong, very strong or stand alone. This truth set resulted in 629 pathogenic and 384 benign variants with *in silico* tool scores available for evaluation.

All variants extracted from the curation databases described above were converted to hg19 genomic coordinates using Mutalyzer (version no. 3)^52^. Data were then uploaded to the dpNSFP (version 4.2)^48^ database in a bed file format.

### Receiver operator curves

The receiver operator curves were generated using the R package; PlotROC^53^. The melt_roc function was used to combine results from each *in silico* tool into single graphs.

### Sensitivity, specificity, positive predictive value and negative predictive value analysis

For each tool, true positive, true negative, false positive and false negative results were determined for each cut-off (table 1). For the assessment of PP3, variants classified as pathogenic were considered as true positives and false negatives if the associated*in silico* tool scores fell above or below the defined cut off respectively. False positives and true negatives were defined as variants with a benign classification and an *in silico* tool score that fell above or below the defined cut off respectively. This assessment was performed at all strength levels provided for each *in silico* tool assessed. For the assessment of BP4, variants classified as benign were considered as true positives and false negatives if the associated *in silico* tool scores fell below or above the defined cut off respectively. False positives and true negatives were defined as variants with a pathogenic classification and an *in silico* tool score that fell below or above the defined cut off respectively.

Sensitivity = true positives/(true positives + false negatives)

Specificity = true negatives/(true negatives + false positives)

PPV = true positives/(true positives + false positives)

NPV = true negatives/(true negatives + false negatives)

Table 1 indicates estimated threshold ranges based on those established by Pejaver et al. for all tools assessed in this study. “[“the range includes the end value; “(“the range excludes the end value. An “-” indicates that the tool was unable to meet the posterior probability threshold. Analyses of the tools BayesDel and VEST4 in the Pajaver et al., 2022 study did not show cut-off scores that reached a posterior probability sufficient for the application of BP4 at a strong level. Given this, these tools were not assessed for BP4 strong level evidence in this study. This was also the case for CADD and PP3 strong level evidence.

### Generation of Positive likelihood ratios

Positive likelihood ratio = Sensitivity / (1 - Specificity)

Approximately 1/3 of tests (37%) were associated with 100% specificity, impacting the ability to calculate meaningful confidence intervals. To address this, confidence intervals were calculated using a bootstrapping method described elsewhere^44^. Briefly, the lowest population specificity that could result in a sample size specificity of 100% is calculated using the formula p = e^1/n^(log 0.5). The average result from 50 tests of 10,000 bootstrap samples drawn from a parametric binomial distribution is then used to estimate the confidence interval. The R package bootLR with the following arguments was used to perform these calculations.

BayesianLR.test(TPo, TPa, TN, TB)

TPo = true positives

TN = true negatives

TPa = total pathogenic

TB = total benign

## Results

The variants included in the analyses presented are represented in Figure 1. This diagrammatic representation demonstrates that the majority of pathogenic variants for *TP53* cluster in the DNA-binding domain, whilst this is not evident for *BRCA1*. Notably for *TERT* pathogenic variants appear to be distributed throughout the gene. In total, 1228 variants (593 pathogenic and 635 benign) in cancer predisposition genes and 1105 variants (721 pathogenic and 384 benign) in a ClinGen curated data set were used as a truth set to assess commonly used *in silico* tools. Results were available for 299, 306, 344, 69, 210 and 1105 variants for *BRCA1, BRCA2, TP53, ATM, TERT* and the ClinGen groups respectively and Figure 1 depicts the location of each of these variants within these genes of interest.

**Figure 1.**
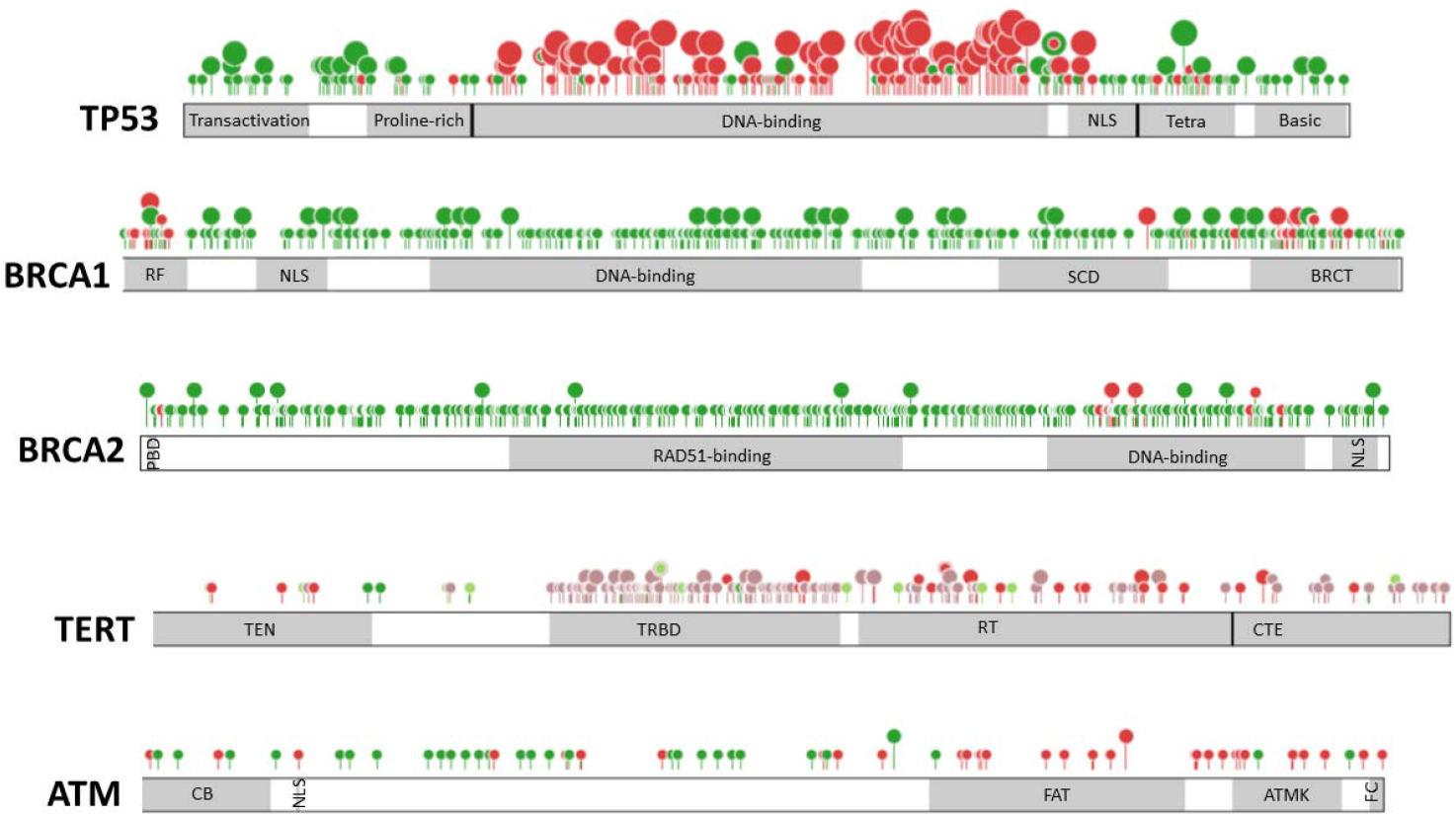
Location of pathogenic and benign variants assessed for *TP53, BRCA1, BRCA2, TERT* and *ATM*. Pathogenic and likely pathogenic variants are represented in red and likely benign and benign variants are represented in green (TERT_1). Variants suspicious for pathogenicity and benignity (TERT_2) are coloured pink and pale green respectively. The number of unique variants reported at a single codon is represented by the size and height of the lollipop symbol (1, 2, 3, 4 or 5 variants). Domain labels are as follows; NLS (nuclear localisation signal), Tetra (tetramerization), RF (ring finger), SCD (serine cluster domain), BRCT (BRCA1 C-terminal), PBD (*PALB2* binding domain, TEN (telomerase essential N-terminal domain), TRBD (telomerase RNA binding domain), RT (reverse transcriptase domain), CTE (C-terminal extension), CB (chromatin binding and substrate interaction), FAT (focal adhesion targeting domain), ATMK (ATM kinase domain), FC (FATC domain). Reference transcripts used are NM_000546, NM_007294, NM_00059, NM_196253 and NM_000051 for *TP53, BRCA1, BRCA2, TERT* and *ATM* plots respectively. Retrieved from ProteinPaint^61^.

### Sensitivity and Specificity

No single *in silico* tool was superior for all five genes reviewed in this analysis (Figure 2). Supporting level, moderate level and strong level evidence cutoffs for each of the *in silico* tools is presented in Table 1. At the PP3 (multiple lines of computational evidence support a deleterious effect on the gene/gene variant) recommended supporting level cut offs, overall, excluding predictions for T*ERT* the tools were found to have a median sensitivity of 78%. Tool performance was poorest for *TERT* predictions with sensitivity of less than 65% for pathogenic *TERT* variants observed for all tools tested. In addition, MutPred predictions for *BRCA1* and Clingen2, CADD predictions for *TP53* and clingen2, VEST4 predictions for *BRCA2* and REVEL, BayesDel and MutPred2 predictions for *ATM* showed a sensitivity of <70%. For the PP3 moderate strength cut offs, only the REVEL predictions for TP53 and BRCA2; the BayesDel predictions for *TP53* and *BRCA1;* and the MutPred2 predictions for *TP53* delivered a sensitivity of greater than 70%. At the level of strong PP3 level of evidence, whilst predictive scores delivered high specificity, they were associated very poor sensitivity at less than 40% with the exception of the BayesDel prediction for *TP53* and the ClinGen truth set (which had sensitivities of 57% and 42% respectively).

**Figure 2.**
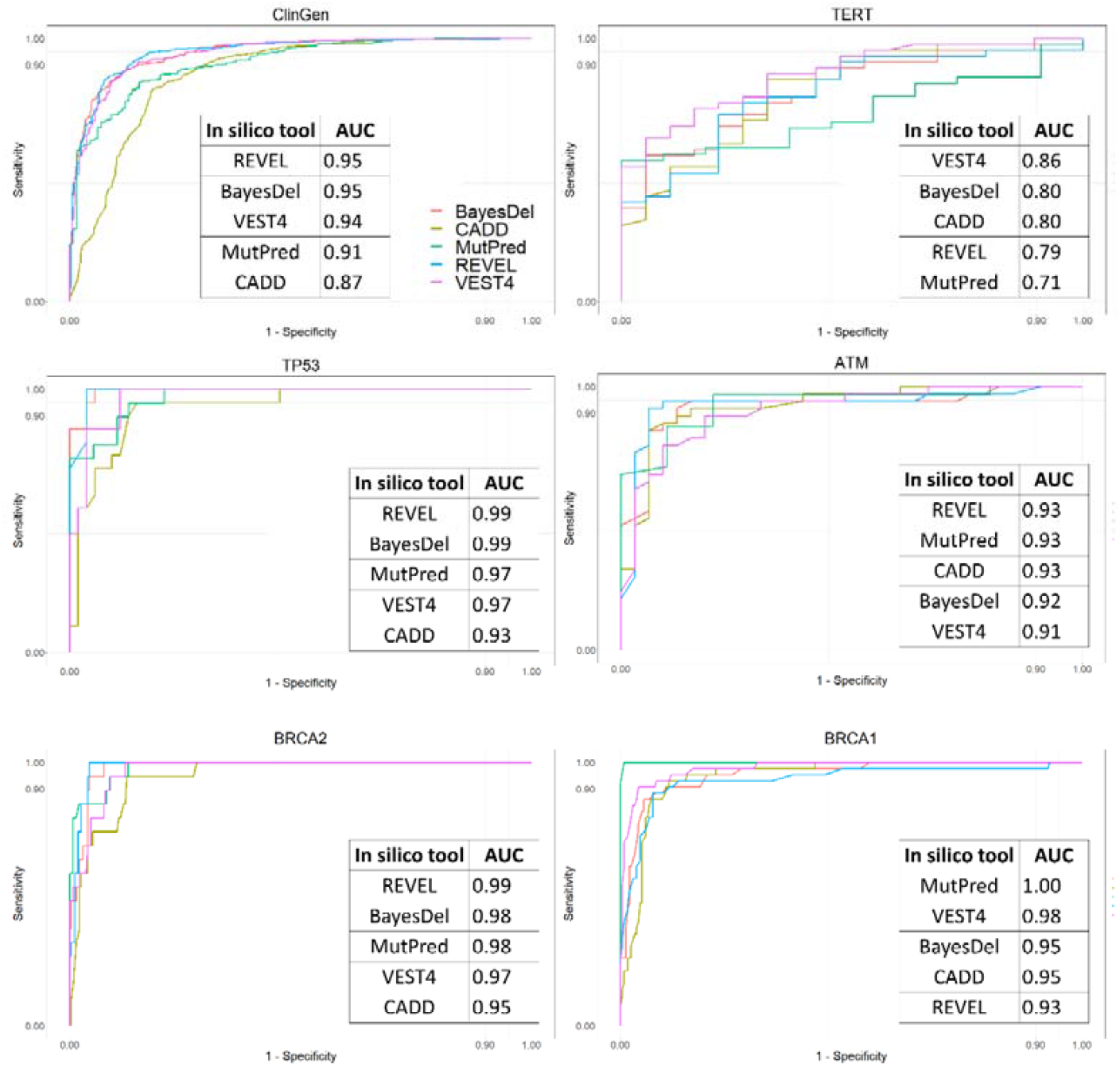
Receiver operator curves (ROC) showing comparative performance of *in silico* predictors for each gene included in this analysis as well as the combined dataset of gene variants curated by ClinGen VCEP committees. Area under the curve (AUC) measurements for each *in silico* tool are annotated for each combination ROC plot arranged in descending order. REVEL scores were associated with the greatest AUC for *BRCA2* and the Clingen curated gene variants. BayesDel scores were associated with the highest AUC for *TP53* (TP53_2) variants. MutPred scores were associated with the highest AUC for *BRCA1* and *ATM* and VEST4 was associated with the highest AUC for *TERT* (TERT_3) variants. ROCs generated using plotROC R package^53^. AUC measurements were calculated using the R package pROC^62^.

Tool specificity at the PP3 supporting level cut off was superior to the sensitivity (median 91%). REVEL and BayesDel predictions for *TP53* and *TERT* were the most inferior (range 67-89%). Specificity was greater than 90% for the majority of predictions at the moderate strength cut offs, the REVEL prediction for *TP53* and the CADD prediction for *BRCA2* being the exceptions. Specificity at the recommended cut-offs for PP3 strong level evidence was very high. Out of the 2113 total variants assessed, false positive calls at the strong level evidence cut-offs were 7 (0.33%), 2 (0.09%), 2 (0.09%) and 1 (0.05%) for BayesDel, REVEL, MutPred2 and VEST4 respectively. BP4 results were further skewed towards high specificity (Figure 3A, Table S9). VEST4 and CADD were associated with the highest sensitivity for BP4 for the cut-offs used.

**Figure 3.**
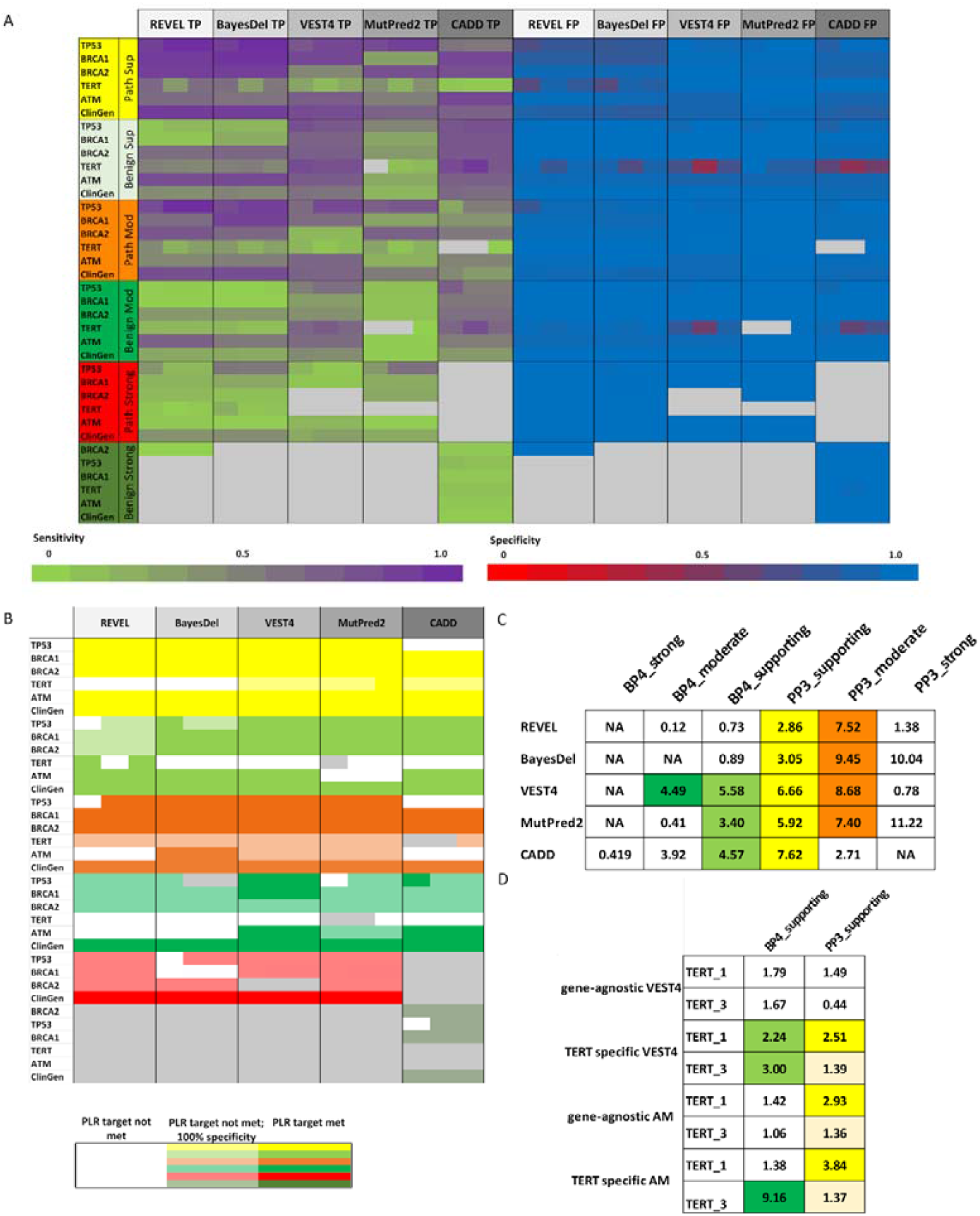
**A**: Heatmap showing comparative true positive (TP) rate and false positive rate (FP) results for the included *in silico* tools at each evidence strength level cut-off score. The rows reflect the truth set tested and the columns reflect the *in silico* tool applied. Where there was more than one truth set used for a gene (such as for TERT), the row was further subdivided into each truth set in numerical order (1, 2, 3) from left to right. Gray cells reflect *in silico* tool predictions that were associated with zero true positive results or *in silico* tool predictions that did not meet posterior probability targets^11^ (see methods). **B**: Heatmap showing comparative positive likelihood ratios (PLRs). The rows are the truth set tested and the columns are the *in silico* tool. White represents *in silico* tool predictions that were associated with less than 100% specificity and a PLR that was below the target PLR for the given strength level. Where there was more than one truth set used for a gene (such as for TERT), the row was further subdivided into each truth set in numerical order from left to right. Three truth sets are included for *TERT* and there are two truth sets for both *TP53* and the ClinGen curated variants. Where there was insufficient data to determine a PLR this is represented in gray. **C**: PLR 95% confidence interval lower limits for the *TP53* gene truth set TP53_2. Where PLR targets are met for PP3_supporting, PP3_moderate, PP3_strong, BP4_supporting, BP4_moderate, BP4_strong this is represented by yellow, orange, red, light green, green and olive green respectively. Where there was insufficient data to determine a PLR this is represented with ‘NA’. Poor sensitivity of BP4 predictions for benign variants have impacted the PLR for REVEL and BayesDel for *TP53* variant predictions.**D**: Comparison of PLRs generated for TERT from gene-agnostic cut-off scores for VEST4 and AlphaMissense versus cut-off scores optimised for improved sensitivity. Bold yellow and green represent PLR 95% CI lower limits that were greater than the target of 2.1 for PP3 supporting and BP4 supporting respectively. Pale yellow reflects a prediction associated with 100% specificity.

### Positive likelihood ratios for PP3

Positive likelihood ratios were determined for all *in silico* tools (Figure 3B). A prediction was considered to have met the target PLR if the lower limit of the 95% confidence interval met or exceeded the target suggested by the ACMG/AMP guidelines (2.1, 4.3, 18.7 for supporting, moderate and strong level evidence respectively). At the cut off scores assessed (Table 1), all tools met or exceeded the target likelihood ratio of 2.1 for PP3 supporting level evidence with the CADD predictions for *TP53* (*TP53_1*) and all predictions for *TERT* being the exceptions. No false positive calls were associated with the VEST4, MutPred2 and CADD supporting level cut offs for *TERT* but poor sensitivity (range 4-62%) resulted in 95% CI lower limits of less than 2.1. For all genes apart from *TERT* tested using the PP3 supporting level cut-offs, MutPred2 predictions were associated with PLRs that met or exceeded the PP3_moderate target of 4.3.

*In silico* predictions less consistently met the PLR targets for PP3 moderate level evidence. The REVEL score predictions for *TP53* (*TP53_1*) failed to meet the PP3_moderate PLR target with a PLR of 3.7 (2.5; 6.19). The VEST4 and REVEL predictions for *ATM* were also slightly inferior to the target for PP3_moderate; 11 (3.658; inf)) and 14.667 (3.875; inf) respectively. There were no false positive calls associated with the REVEL, BayesDel, MutPred2 and VEST4 predictions for *TERT* (TERT_1 and TERT_2); however, the sensitivity for pathogenic variants was poor thus the lower 95%CI limits were all were less than 4.3. CADD performed the worst with only the predictions for *BRCA1* and *BRCA2* exceeding the 4.3 target.

While all PLRs exceeded the target for the cut-offs recommended for PP3_strong, poor sensitivity resulted in the lower limit of the 95%CIs falling below the target of 18.1 for *TP53, BRCA1* and *BRCA2. TERT* and *ATM* had insufficient numbers of variants tested to assess a PLR target of 18.1. Use of CADD scores at a strong strength level has not been determined.

### Positive likelihood ratios for BP4

To enable comparison, BP4 results have been determined using PLRs. Poor sensitivity resulted in *in silico* tools inconsistently meeting target PLRs for BP4 (Figure 3B, 3C). Apart from predictions for *TERT* variants, the tools were however highly specific. The best performing tool was VEST4. All genes tested met the target PLRs for the VEST4 cut-offs for supporting and moderate level evidence apart from *TERT*. The lower limit of the 95% CI was also lower than 4.1 for *BRCA2* for the BP4 moderate level evidence cut-off; however, no false positive results were seen and further, at the cut-off for supporting level evidence, the lower limit of the 95% CI was 5.43, exceeding the moderate level target PLR.

At the cut offs for BP4 strong level, only CADD predictions for *BRCA1, BRCA2* and the ClinGen groups were associated with true positive results. No false positive results were seen associated with the strong level cut-off scores for CADD; however poor sensitivity resulted in all 95%CIs dropping below the target of 18.7.

### Optimisation of *in silico* cut off scores for TERT

In light of the suboptimal performance of gene-agnostic *in silico* score cut offs for TERT, the ROC curves were reviewed to determine if the PLRs could be optimised. Of the tools reviewed, VEST4 was associated with the highest AUC; 0.86 (Figure 2). Sensitivity and specificity calculations were suggestive of this being predominantly due to superior specificity. Therefore, the VEST4 cut-off scores for PP3 supporting and BP4 were revised down and up respectively to improve sensitivity. Revising down the VEST4 cut off score for PP3 supporting to 0.5 and above, the sensitivity improved to 67% and 59% for TERT_3 and TERT_1 respectively. Specificity improved to 0.9 and 1 for TERT_3 and TERT_1 respectively. The PLR 95%CI lower limit slightly improved to 2.5 and 1.4 for TERT_3 and TERT_1 respectively (Figure 2D). For BP4, a VEST4 score of equal to or below 0.16 was associated with improved sensitivity to 50% and 50% respectively and specificity of 90% and 97% respectively. The PLR 95% CI lower limit was 2.2 and 2.9 for TERT_3 and TERT_1 respectively (Figure 2D). While the gene-agnostic predictions of pathogenicity and benignity generated by the newly released AlphaMissense^41^ *in silico* tool show similarly poor PP3 sensitivity (50%) and BP4 specificity (61%) (TERT_1 dataset), a similar cut off adjustment also significantly improved the predictive accuracy of the tool. Applying a pathogenic prediction at a cut off score of greater than 0.40 and a benign prediction at a cut off score of less than 0.09 was associated with a PP3 sensitivity for pathogenic variants to 59% and a BP4 specificity for benign variants of 100% respectively. PLRs also improved as shown in Figure 2D.

### MISCAST prediction gene conformance to missense variant-protein structural and functional impact relationships identified in large multi gene training sets

*In silico* tools use ‘black-box’ algorithms and are frequently composites of other online*in silico* tools and the weighting to each source tool to deliver a ‘score’ for a given variant is opaque. To investigate how protein structural and functional impact predictions may be influencing *in silico* scores for the gene variants included in this study, the pathogenic 3D Feature index (P3DFi)^20^ was compared with the previously examined tools. The MISCAST algorithm mines and annotates amino acid residues for structural, physiochemical and functional features to deliver a P3DFi score. Similar protein feature databases, such as UniProt^37^, protein-protein interaction databases (PDBsum^54^) and protein secondary structure databases (DSSP^55^), have also been incorporated into MutPred2^14^, PolyPhen2^56^ and Mutation Assessor^57^ (and therefore REVEL^13^, BayesDel^15^ and CADD^18^). In addition, VEST4^16^ incorporates protein feature data from these data sources via SNVBox^40^. P3DFi scores for *BRCA1, BRCA2* and *ATM* showed similar enrichment for pathogenic variants as the MISCAST gene variant dataset^20^ (Figure 4). Lower P3DFi scores (<-1) appeared, however, to be more enriched for pathogenic *TERT* variants compared to *TP53, BRCA1, BRCA2, ATM* and the complete MISCAST dataset of variants from 1,330 disease-causing genes (Figure 3, Tables S10, S11, S12, S13, S14). Scores -1 to 2 were more enriched for benign *TP53* variants compared to the other groups which may account for the poor sensitivity of BP4 predictions for benign and poor specificity of PP3 predictions for pathogenic *TP53* variants respectively.

**Figure 4.**
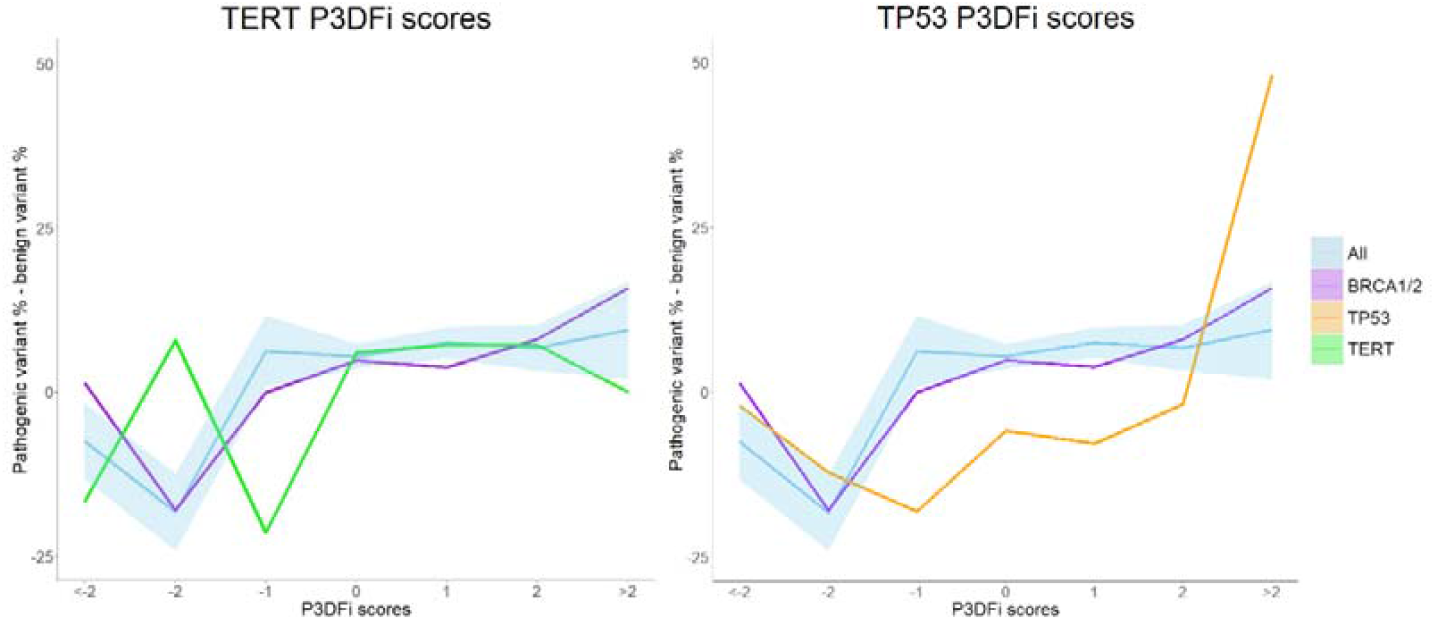
The relative enrichment of pathogenic variants at P3DF1 scores ranging from <-2 to >2 for the genes included in this study as well as the MISCAST combined dataset of 1,330 disease causing genes^20^. Relative enrichment of pathogenic variants is determined by the percentage of pathogenic variants minus the percentage of benign variants at each P3DFi score. Increasing P3DF1 score is associated with increasing predicted structural impact from a given missense variant. P3DFi scores of *TERT* (TERT_3) and *TP53* (TP53_2) are shown in orange and coral respectively. Comparative P3DFi scores for the MISCAST combined dataset of 1,330 disease causing genes are shown in blue. The standard error of the mean pathogenic variant enrichment for *ATM, BRCA1, BRCA2* and the combined MISCAST data set of 1,330 disease causing genes is shown by the shaded pale blue region. The mean enrichment of pathogenic variants at each P3DFi score for a combined dataset of BRCA1 and BRCA2 variants are shown is shown in purple. Plots generated using gglot2; https://ggplot2.tidyverse.org.

### Evidence stacking using SpliceAI predictions

Most *in silico* tools do not consider the potential impact of missense variants on splicing. To account for the possibility of cryptic splice site creation by missense variants, *in silico* splice impact tool predictions are often incorporated into PP3/BP4 use guidelines^58^. A SpliceAI cut off score of > 0.38 (found previously to be associated with a 90% sensitivity for impact on splicing^1^) as an additional criterion to meet PP3_supporting slightly increased the sensitivity of PP3 supporting level evidence for variants with a classification of pathogenic compared with use of REVEL, VEST4, MutPred2, CADD or BayesDel alone (Table 2, Table S15, S16). One further false positive result (*RYR1:c*. 9353C>T) would result from applying PP3_supporting for a SpliceAI delta change prediction of >0.38. A SpliceAI delta cut-off score of < 0.2 as an additional requirement for the application of BP4 did not address any false positive calls of BP4 to variants classified as pathogenic. It did, however, reduce the number of variants with a benign classification meeting the BP4 criterion, further reducing sensitivity.

**Table 2.**
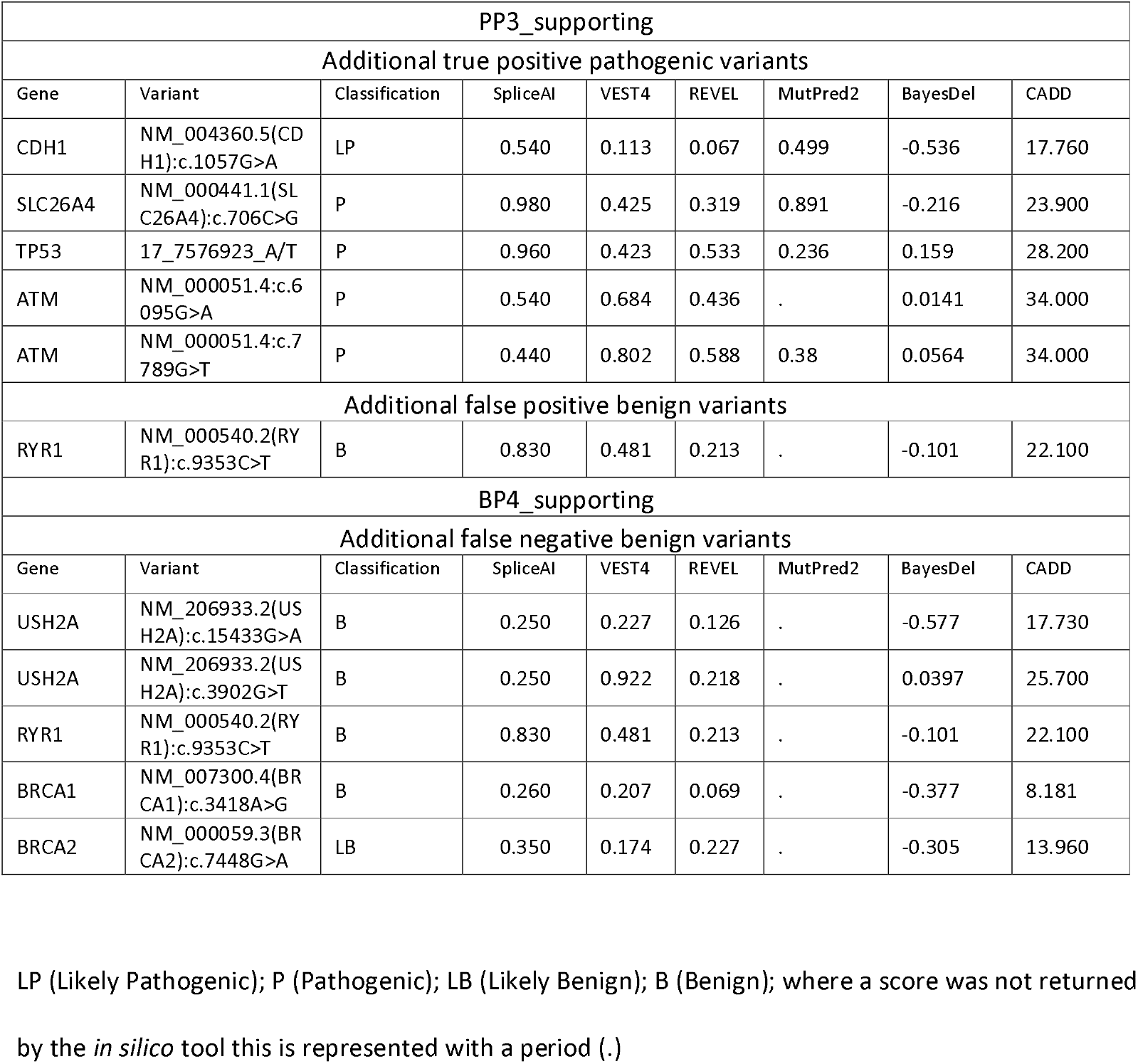
Variants with a SpliceAI score that altered the application of PP3 or BP4 criteria.

## Discussion

### Performance of in silico tools

Where possible, the ACMG/AMP guidelines^9^ recommend statistical analysis of curation rules to assess if their application will result in final variant classifications with high posterior probabilities of being correct. Likely pathogenic classifications should be associated with a posterior probability of at least 0.9 and pathogenic classifications should have a posterior probability of >0.99^22^. Using the current approaches for combining evidence using the ACMG/AMP guidelines, a target posterior probability can then be determined for each strength level of an individual rule (supporting, moderate, strong and very strong). By sorting the scores for a combined dataset of pathogenic and benign variants from ClinVar and gnomAD, Pejaver et al., 2022^11^ were able to determine score intervals for a given *in silico* tool that would result in these predefined posterior probability targets.

At the interval cut-offs suggested by that work, in general the *in silico* tools assessed in this study demonstrated high specificity at the expense of sensitivity. Where tools did not meet the target PLRs this was most commonly due to poor sensitivity. In the context of diagnostic curation, a trade-off in favour of high specificity is appropriate as high confidence in application of PP3 or BP4 is preferable. With the exception of BP4 predictions for *TERT* and PP3 predictions for *TERT* and *TP53*, the results from this study demonstrate the gene-agnostic use of PP3 and BS4 at multiple strength levels is associated with acceptable specificity (at least 80% at supporting level and at least than 90% at moderate level). This supports the use of this approach when there are insufficient variants with established non-VUS classifications to perform a gene-specific validation. These results also highlight the importance of considering specificity and sensitivity in combination with the PLR when assessing the strength of evidence a tool or ACMG/AMP rule provides. Relying on the PLR results alone may result in rejection of an *in silico* tool that is highly specific if the sensitivity is low.

These results also show that *in silico* prediction tool performance is variable. For example, *in silico TP53* BP4 and PP3 predictions were associated with poor sensitivity and poor specificity respectively.

The associated P3DFi scores are suggestive that the *TP53* protein may have increased tolerance for physicochemical changes associated with missense variants compared to other disease-causing genes commonly included in training datasets. This is further supported by a lack of constraint for missense variants in *TP53* even within the DNA binding domain^59,60^. *In silico* predictions for*TERT* variants were the most inferior. The poor concordance seen in all *TERT* variant datasets appeared to be driven by poor sensitivity for pathogenic variants and poor specificity for benign variants. *TERT* is a highly constrained gene with less enrichment of benign variants seen in P3DFi scores < -1. As *in silico* tools are trained using datasets of large numbers of genes, scores may be insensitive to genes encoding proteins that are highly sensitive to small physicochemical changes. Revising cut off scores for the *in silico* predictor with the highest AUC (VEST4) to improve sensitivity was associated with improved PLRs for both PP3 and BP4. Similar improvements in PP3 sensitivity and BP4 specificity were also seen when AlphaMissense cut off scores were revised in the same direction. These results support the use of gene-specific *in silico* tool validation where there are sufficient data to do so. Selecting different tools and cut offs for different genes however, needs to be weighed up against the problems this creates for variant filtering in large gene panels and exomes prior to manual curation. While no one tool performed the best for all genes studied in this analysis (Figure 2) a commonly used tool associated with adequate PLRs or 100% specificity following a gene-specific validation may be appropriate for use if this simplifies bioinformatic workflows.

### The use of truth sets of variants with standardized curation

*In silico* tools use a range of data associated with training sets of variants such as population variation, evolutionary conservation, physiochemical differences between reference and variant amino acids and predicted disruptions to the 3D protein structure to predict the impact of variants on protein function. The most commonly used training sets include the ClinVar, HGMD and gnomAD databases. These attribute-pathogenicity associations are therefore determined using sets of highly heterogeneous genes. These associations may not necessarily be generalisable to a given individual gene. The ClinVar and HGMD databases collate curations from submissions from individual diagnostic laboratories. The given curations are not standardized or vetted and may be influenced by the phenotype of the individual harbouring that variant. For example, a pathogenic variant causative of a disease not associated with the particular presenting phenotype may be reported as non-pathogenic. Minimal data are available regarding the evidence used for most individual curations, including the use of*in silico* tool evidence. Increasing efforts, by groups such as ClinGen have been under way to standardise curations^1,10^. Where possible this study has utilised these guidelines to create truth sets of variants curated according to standardised approaches. This has provided the best available data to benchmark the *in silico* tools assessed in this validation study.

### Addition of SpliceAI predictions

REVEL, VEST4, BayesDel and MutPred2 do not incorporate predictions of potential splicing disruption caused by missense variants. Whilst CADD is influenced by the tendency of variants that impact splicing to be depleted it is also not designed with a focus on splicing predicton^17,18^. SpliceAI delta scores of greater than 0.38 are associated with a 90% sensitivity for splicing alterations as characterized by RNA studies^1,58^. While RNA data was not assessed in this study, the use of a delta score cut off of 0.38 or greater to apply PP3_supporting in addition to another *in silico* tool did result in increased true positive results with only one additional false positive result. The requirement of a delta score of 0.2 or less has also been incorporated into ClinGen guidelines for the application of BP4. For the variants assessed in this study, the use of this cut-off did not prevent any false positive calls of BP4 for a pathogenic variant. It did, however, result in higher false negative calls.

## Limitations

Where possible, truth sets that contained curations with multiple applied lines of validated evidence were used in this analysis. The ClinGen, *BRCA1* and *BRCA2* curation databases provided standardized curations with detailed information on the use of evidence. *TERT* variants were also curated using a standardized approach in line with current recommendations^2,8^. Unfortunately, such a resource was not available for *ATM;* while the variants included in the LOVD3 database are collated by a single curator, complete details on the curation process are not publicly available. Where possible, truth sets were limited to variants that met a non-VUS classification without the use of PP3 or BP4. This was possible for *TERT, TP53* and the ClinGen gene variant data. The dependency on*in silico* data for the classification of *BRCA1, BRCA2* and *ATM* variants could not be determined from the publicly available data. The inclusion of variants with classifications that were dependent on results from the *in silico* tools assessed in this study may have resulted in an overestimation of the accuracy of the tools for these genes. Apart from CADD^18^, which was not trained on datasets of curated variants, the variants in this study may also have been included in the original training datasets for the *in silico* tools assessed. This may also have overestimated the accuracy of these *in silico* tools. Given, however, the training datasets included large numbers of different genes, it’s unlikely the underlying algorithms would be strongly influenced by the individual genes assessed in this study.

## Conclusions

The currently available in silico tools are trained on truth sets comprised of variants from multiple genes. The function of proteins such as TERT may be impacted by missense variants by mechanisms that are under-represented in these combined datasets. The results from this study highlight the importance of gene-specific validation of in silico tool use especially if PP3/BP4 is applied at moderate or strong levels. We have also identified that while *in silico* tool predictions are associated with high specificity, improving tool sensitivity remains an area of need. The addition of a splicing impact *in silico* tool such as SpliceAI may improve sensitivity for pathogenic variants.

## Data Availability

All data produced in the present work are contained in the manuscript.

## Data and code availability

Additional data supporting the findings of this study are available in either the manuscript and/or supplemental material.

## Acknowledgments

N.N. is supported by an Annie Bishop doctoral degree scholarship in cancer research. K.F. is supported by the Alex Gadomski Fellowship, funded by Maddie Riewoldt’s Vision, and an NHMRC grant (GNT 2020517). J.D. is supported by a Select Foundation Fellowship. T.M.B. was supported by The Arcus Foundation (Australia), the Neil and Norma Hill Foundation, and Luminesce Alliance - Innovation for Children’s Health, a not for profit cooperative joint venture between the Sydney Children’s Hospitals Network, Children’s Medical Research Institute, the Children’s Cancer Institute, the University of Sydney and the University of New South Wales, established with the support of the NSW Government.

## Author Information

Conceptualization: N.N., S.L., J.D.; Data curation: N.N., A.A., T.B.; Formal analysis: N.N., A.A., T.B.; Investigation: N.N., A.A., T.B.; Methodology: N.N., S.L.; Resources: A.A., T.B; Software: K.F.; Supervision: J.D., K.F., S.L.; Validation: N.N., J.D., T.B.; Visualization: N.N., K.F; Writing-original draft: N.N.; Writing-review & editing: N.N., T.B., K.F., S.L., J.D.

## Ethics Declaration

All genetic data analyzed and reported in this study have been previously published, are discussed in articles that are in press and/or stem from publicly available databases such as ClinVar (https://www.ncbi.nlm.nih.gov/clinvar/). No institutional review board or research ethics committee approval was required.

## Conflict of Interest

All authors declare no conflicts of interest.

## Additional Information

The online version of this article contains supplemental material, which is available to authorized users.

## Web Resources

ClinGen Evidence Repository; https://erepo.clinicalgenome.org/evrepo/

*BRCA1* and *BRCA2* Mutation Databases (University of Utah); https://arup.utah.edu/database/BRCA/

*ATM* gene home page; Lieden open variation database (LOVD3); https://databases.lovd.nl/shared/genes/ATM

dbNSFP database; https://sites.google.com/site/jpopgen/dbNSFPgnomADbrowser; https://gnomad.broadinstitute.org/

ClinVar; https://www.ncbi.nlm.nih.gov/clinvar/

Mastermind Genomic Search Engine; https://mastermind.genomenon.com/

Variant Effect Predictor; http://grch37.ensembl.org/Homo_sapiens/Tools/VEP

MIssense variant to protein StruCture Analysis web SuiTe; https://miscast.broadinstitute.org/

ProteinPaint; https://proteinpaint.stjude.org/

Ggplot2; https://ggplot2.tidyverse.org/

## Notes

### Competing Interest Statement

The authors have declared no competing interest.

### Author Declarations

The study used only openly available human data that were originally located at multiple databases, all referenced in the body of the manuscript. Please see manuscript for more details.

